# The impact of post-hospital remote monitoring of COVID-19 patients using pulse oximetry: a national observational study using hospital activity data

**DOI:** 10.1101/2022.01.11.22269068

**Authors:** Theo Georghiou, Chris Sherlaw-Johnson, Efthalia Massou, Stephen Morris, Nadia E Crellin, Lauren Herlitz, Manbinder S Sidhu, Sonila M Tomini, Cecilia Vindrola-Padros, Holly Walton, Naomi J Fulop

**Affiliations:** Nuffield Trust, 59 New Cavendish St, London W1G 7LP; Department of Public Health & Primary Care, University of Cambridge, Cambridge, UK; Department of Applied Health Research, University College London, Gower Street London, WC1E 6BT, UK; Health Services Management Centre, School of Social Policy, College of Social Sciences, University of Birmingham, 40 Edgbaston Park Rd, Birmingham, B15 2RT; Department of Targeted Intervention, University College London, Charles Bell House, 43-45 Foley Street, London, W1W 7TY

## Abstract

**Background:** There was a national roll out of ‘COVID Virtual Wards’ (CVW) during England’s second COVID-19 wave (Autumn 2020 – Spring 2021). These services used remote pulse oximetry monitoring for COVID-19 patients following discharge from hospital. A key aim was to enable rapid detection of patient deterioration. It was anticipated that the services would support early discharge and avoid readmissions, reducing pressure on beds. This study is an evaluation of the impact of the CVW services on hospital activity.

**Methods:** Using retrospective patient-level hospital admissions data, we built multivariate models to analyse the relationship between the implementation of CVW services and hospital activity outcomes: length of COVID-19 related stays and subsequent COVID-19 readmissions within 28 days. We used data from more than 98% of recorded COVID-19 hospital stays in England, where the patient was discharged alive between mid-August 2020 and late February 2021.

**Findings:** We found a longer length of stay for COVID-19 patients discharged from hospitals where a CVW was available, when compared to patients discharged from hospitals where there was no CVW (adjusted IRR 1·05, 95% CI 1·01 to 1·09). We found no evidence of a relationship between the availability of CVW and subsequent rates of readmission for COVID-19 (adjusted OR 0·95, 95% CI 0·89 to 1·02).

**Interpretation:** We found no evidence of early discharges or reduced readmissions associated with the roll out of COVID Virtual Wards across England. Our analysis made pragmatic use of national-scale hospital data, but it is possible that a lack of specific data (for example, on which patients were enrolled) may have meant that true impacts, especially at a local level, were not ultimately discernible.

**Funding:** This is independent research funded by the National Institute for Health Research, Health Services & Delivery Research programme and NHSEI.

**Research in context:** *Evidence before this study:* Post-hospital virtual wards have been found to have a positive impact on patient outcomes when focussed on patients with specific diseases, for example those with heart disease. There has been less evidence of impact for more heterogenous groups of patients. While these services have been rolled out at scale in England, there has been little evidence thus far that post-hospital virtual wards (using pulse oximetry monitoring) have helped to reduce the length of stay of hospitalised COVID-19 patients, or rates of subsequent readmissions for COVID-19.

*Added value of this study:* This national-scale study provides evidence that the rollout of post-hospital discharge virtual ward services for COVID-19 patients in England did not reduce lengths of stay in hospital, or rates of readmission.

*Implications of all the available evidence:* While there is currently an absence of evidence of positive impacts for COVID-19 patients discharged to a virtual ward, our study emphasises the need for quality data to be collected as part of future service implementation.

## Introduction

During the early months of the COVID-19 pandemic in England, a number of remote home-based monitoring programmes were set up with the aim of using pulse oximetry to detect early signs of deterioration in people with COVID-19. Managed by primary and secondary care providers, the services aimed to safely manage patients - identified from within the community, from Emergency Departments (ED), or following hospital discharge – outside of hospital, to provide reassurance and rapid escalation where necessary.^1–7^ Similar remote home monitoring programmes were also developed in several other countries.^8^

During England’s second COVID-19 wave in November 2020, the NHS in England launched a national roll out of community-based home monitoring services under the banner of ‘COVID Oximetry @ home’ (CO@h).^9^ In January 2021, there followed a national launch of post-hospital discharge ‘COVID Virtual Ward’ (CVW) models.^10^ CO@h models (generally managed by primary care services) were intended to focus on lower-acuity (less severely ill) COVID-19 patients diagnosed in the community, with the aims of self-escalation and more appropriate admission, while CVW models (generally managed by secondary care) were intended to support the early discharge of hospitalised higher-acuity patients and prevent unnecessary readmissions.

Prior to COVID-19, the NHS in England made clear its ambition to develop remote monitoring services to better support people with long-term conditions out of hospital.^11^ Virtual ward models have been proposed as one way of easing the transition of patients being discharged home, and there is some evidence that such models (distinguished from less intensive telemonitoring) may help to reduce mortality and readmissions, especially where the intervention is focussed on patients with a specific disease, for example heart failure.^12^ We discuss the impact of the implementation of CO@h models elsewhere^13^; the focus of this study is the impact of post-hospital discharge CVW models.

National guidance set out the recommended protocol for new CVW services.^14^ It was to be available for adults in hospital with a primary diagnosis of COVID-19, where clinical assessments had determined that the patient was suitable for discharge. Specifically, suitable patients were likely to be those with oxygen saturations of 95-100%, ‘NEWS2 scores’ less than 3 (this is an index based on physiological readings, linked to risk of clinical deterioration)^15^ and improving clinical trajectories. Patients with saturation levels of 93-94% and with improving trajectories (symptoms, signs, blood results and chest X-rays) might also be considered for the service.

Enrolled patients were to be given a pulse oximeter (with instructions), and have an agreed discharge and escalation plan. This would include monitoring arrangements, a diary for use by the patient, and a hospital number to call for advice or escalation. The patient would be asked to take three readings per day following discharge, and call the service (or other available emergency services, if out of hours) immediately if a reading was less than 92%. The patient was also to be proactively contacted by phone every day, as if still part of a hospital-based ward round. By day 14 on the CVW, or earlier where clinically appropriate, the patient would be discharged from the service, and the pulse oximeter returned for reuse.^14^

However, the national roll out of CVW was not intended to supplant existing services, and nor was it intended to be entirely prescriptive. Our related study of a number of such services in England demonstrated considerable variability in implementation models, including in patient eligibility criteria, onboarding processes and monitoring processes (including the use of digital technologies and frequency of contact).^16^ A number of the services were managed as joint integrated community (pre-hospital) and hospital (post-discharge) models.

In facilitating the national rollout of services, NHS England stated that “CVWs have been proven to reduce admissions/bed occupancy and improve length of stay”, and so could act to mitigate the pressure on hospital beds, which was severe at the time.^17^ However, evidence of the impact on hospital activity has been lacking. A US study found a reduction in rates of ED presentation or readmission for a group of 225 patients enrolled onto a post-discharge monitoring programme,^18^ and two studies (from the Netherlands, and a case study from England) estimated reduced lengths of hospital stays, but with small patient cohorts and uncontrolled methods.^19,6^ Elsewhere, where outcomes have been reported, these have tended to focus on rates of patient escalation (including readmissions) in the absence of a comparator. Occasionally these are considered against a perceived optimal level (for example, a 10% conversion rate to admissions for ambulatory care pathways^2^), or the reported performance of services elsewhere.^1–4,7,20,21^

Data on which patients were enrolled to CVWs in England was not collected, but information was assembled on launch dates of CVW at each hospital trust. By late February 2021 the large majority of acute hospital trusts in England had a service available for use. Our analysis aimed to use patient-level admissions data to provide quantitative evidence of the impact of the service, to contribute to future development of post-discharge home monitoring programmes during the remainder of the COVID-19 pandemic, and beyond.

The primary aim of this paper was to address the following research question:

What was the impact of COVID Virtual Wards on the length of stay of COVID-19 hospitalisations, and rates of subsequent readmission for COVID-19?

A secondary aim, in the context of COVID-19’s widely reported differential effects on outcomes for groups of patients (for example, by gender, age and ethnicity),^22,23^ was to reflect on associations found between patient characteristics and hospital outcomes.

## Methods

### Data sources

The data sets used in this analysis are provided in Table 1.

**Table 1.**
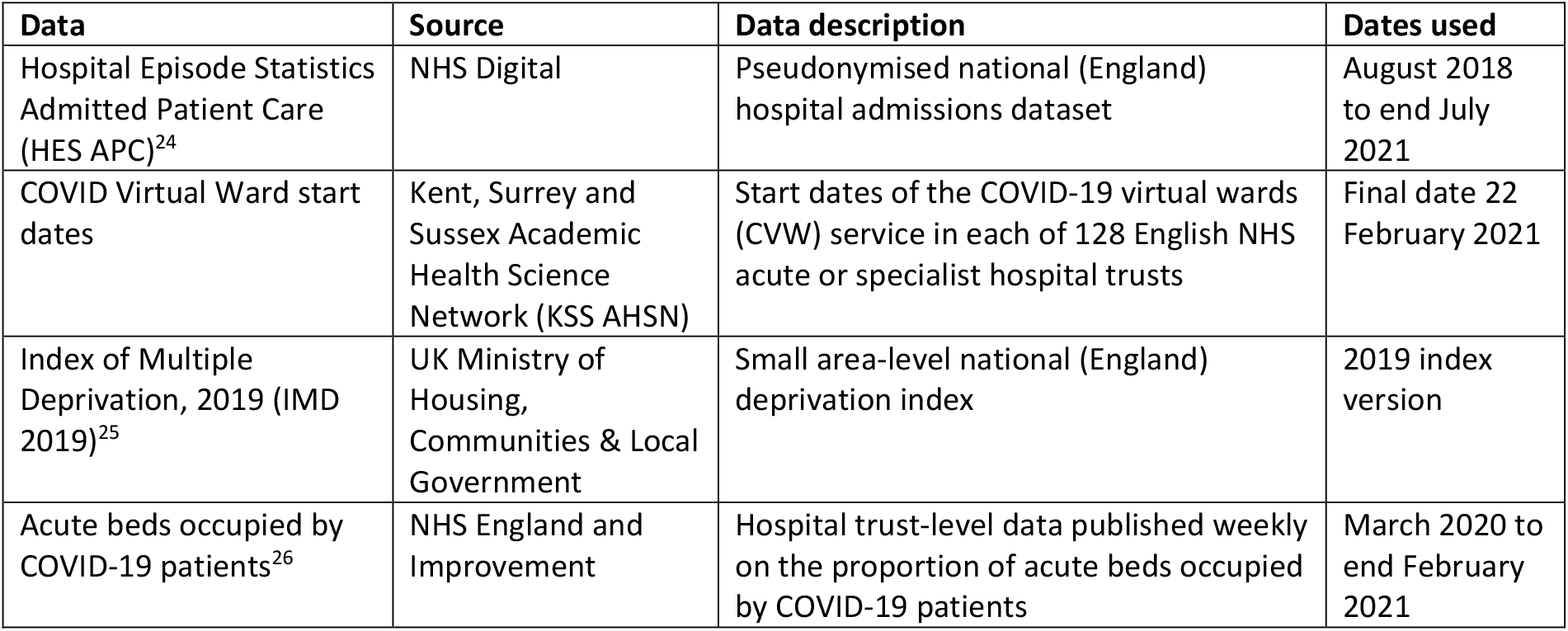
Data sets used in analysis.

| **Data** | **Source** | **Data description** | **Dates used** |
| --- | --- | --- | --- |
| Hospital Episode Statistics Admitted Patient Care (HES APC)<sup>24</sup> | NHS Digital | Pseudonymised national (England) hospital admissions dataset | August 2018 to end July 2021 |
| COVID Virtual Ward start dates | Kent, Surrey and Sussex Academic Health Science Network (KSS AHSN) | Start dates of the COVID-19 virtual wards (CVW) service in each of 128 English NHS acute or specialist hospital trusts | Final date 22 February 2021 |
| Index of Multiple Deprivation, 2019 (IMD 2019)<sup>25</sup> | UK Ministry of Housing, Communities & Local Government | Small area-level national (England) deprivation index | 2019 index version |
| Acute beds occupied by COVID-19 patients<sup>26</sup> | NHS England and Improvement | Hospital trust-level data published weekly on the proportion of acute beds occupied by COVID-19 patients | March 2020 to end February 2021 |

### Setting and participants

From HES APC data, we extracted information on all individuals discharged alive from 123 English hospital trusts where there had been a confirmed or suspected COVID-19 ICD-10 diagnosis code (U071 or U072) recorded as a primary diagnosis at any point during the inpatient stay. We included all patients discharged between 17 August 2020 and 28 February 2021, a period covering the beginning, the peak, and the start of the decline of England’s second COVID-19 wave.^27^ Where a patient had two or more relevant inpatient stays, all stays were included in our analysis. The 123 acute trusts were selected from the KSS AHSN list of 128, having excluded four specialist trusts (not expected to treat patients with COVID-19 as a primary cause of the admission), and one non-specialist acute trust whose CVW start date was not known. We additionally extracted limited data on numbers of similar COVID-19 discharges for all trusts in the HES APC data, to compare the number in our analysis with national counts of similar patients.

### Analytical approach

We developed multivariate models to examine the impact of the availability of CVW (the primary independent variable of interest) on two outcomes: the length of stay (LOS) of the COVID-19 inpatient stay, and on subsequent readmissions for COVID-19.

### Variables

In our analyses we included a range of factors likely to be associated with LOS and rates of readmissions for COVID-19 patients;^28–33^ see Table 2. Time period categories were included to take account of fluctuating baseline LOS and rates of readmissions over the analysis period.^34^ The proportion of beds occupied by COVID-19 patients was included to represent COVID-related bed pressures.

**Table 2.** Factors included in models.

| **Modelling Factors** | **Categories assigned to each COVID-19 patient discharge** |
| --- | --- |
| Age at admission | 0-17, 18-49, 50-64, 65-79, 80s+ |
| Gender | Male, Female |
| Ethnic group | Asian, Black, White, Mixed, Other, Unknown |
| Charlson comorbidity index category<sup>35</sup> | 0, 1, 2, 3, 4, 5, 6+ |
| Whether the stay was the person's first COVID-19 hospital stay | Yes, No |
| Whether the inpatient stay was an emergency admission | Yes, No (elective) |
| Deprivation quintile Index of Multiple Deprivation (2019) | 1 – most deprived to 5 – least deprived, with a sixth category for unknown area |
| Time period of the discharge date | 14 categories, each covering a 14-day period starting from 17 August 2020 |
| Proportion of all acute beds occupied by COVID-19 patients | A trust- and week-specific continuous measure (mapped to week of discharge) |

Age, gender, ethnic group, hospital trust, emergency admission information, time period of discharge were all taken from information recorded against the inpatient COVID-19 stay itself, as was the lower super output area (LSOA) of residence of the patient, which was used to add the deprivation quintile. The Charlson comorbidity category index score^35^ was calculated using HES APC data (specifically, diagnostic information) from two years (730 days) prior to the COVID-19 admission date. The proportion of acute beds occupied by COVID-19 patients was assigned to the appropriate week of the date of discharge.

From the COVID-19 inpatient stay, we recorded the LOS as the discharge date minus the admission date. In our analyses, we replaced all LOS of greater than 60 days with 60 days to reduce the potentially distorting impact of very long lengths of stay.

Using subsequent HES APC data, we recorded the occurrence of any readmission for COVID-19, to any hospital, within 28 days of the COVID-19 stay discharge date. Here we included any confirmed or suspected COVID-19 ICD-10 diagnosis code, recorded as either a primary or secondary diagnosis on the admission episode of the inpatient stay.

The variable indicating the availability of COVID-19 virtual ward was assigned depending on the hospital trust and date: for any individual trust, every discharge from the day of the CVW service start date onwards was assigned as having a CVW available, while all discharges before that date were assigned to having no CVW available. Where a trust was known to have not implemented a CVW by the end of the analysis period, all that trust’s discharges were assigned as having no CVW available.

### Statistical analysis

Basic descriptive statistics were initially used to provide information on the characteristics of all patients included in the analysis, and also split into two mutually exclusive groups: COVID-19 patients discharged from a hospital trust where a CVW was, and was not, available. We compared differences between the groups using Pearson’s chi-squared test for categorical variables and a two independent samples t-test for one continuous variable.

We calculated unadjusted means of COVID-19 LOS and rates of COVID-19 readmission for all categories of patient characteristics. Negative binomial regression was used to examine the relationship between independent variables and LOS,^36^ and logistic regression was similarly used for occurrence of any readmission. To account for clustering at the level of the hospital trust, generalised estimating equation (GEE) approaches were used.

To investigate the robustness of our findings, we carried out a number of sensitivity analyses. For both outcomes, we tested two alternatives for the time period of discharge variable: 7 days and 28 days, and also tested including data from England’s first COVID-19 wave (specifically from 2 March 2020). We also tested two further LOS outcomes: one untrimmed at 60 days (that is, the crude LOS, however long), and another where we disregarded episodes of care at the beginning of the inpatient stay, where these appeared to predate the COVID-19 diagnosis. Moreover, we iteratively examined the statistical significance of each independent variable as well as the impact of their order, by constructing our models step-by-step.

All analyses were carried out in SAS v9·4 (SAS Institute, North Carolina, US).

### Ethical considerations

The use of HES APC data was governed by a data sharing agreement with NHS Digital covering NIHR RSET analysis (DARS-NIC-194629-S4F9X). A protocol covering this analysis (as one part of a wider study) received ethical approval from the University of Birmingham Humanities and Social Sciences ethics committee (ERN_13-1085AP39) and was categorised as a service evaluation by the HRA decision tool and UCL/UCLH Joint Research Office (Jan 2021).

## Results

### Participants

We found 139,619 discharges between 17 August 2020 and 28 February 2021 where the patient had a primary diagnosis of COVID-19 recorded during their inpatient stay, and had not died in hospital. These discharges belonged to 129,461 individuals. Table 3 shows characteristics of these COVID-19 patient discharges.

**Table 3.**
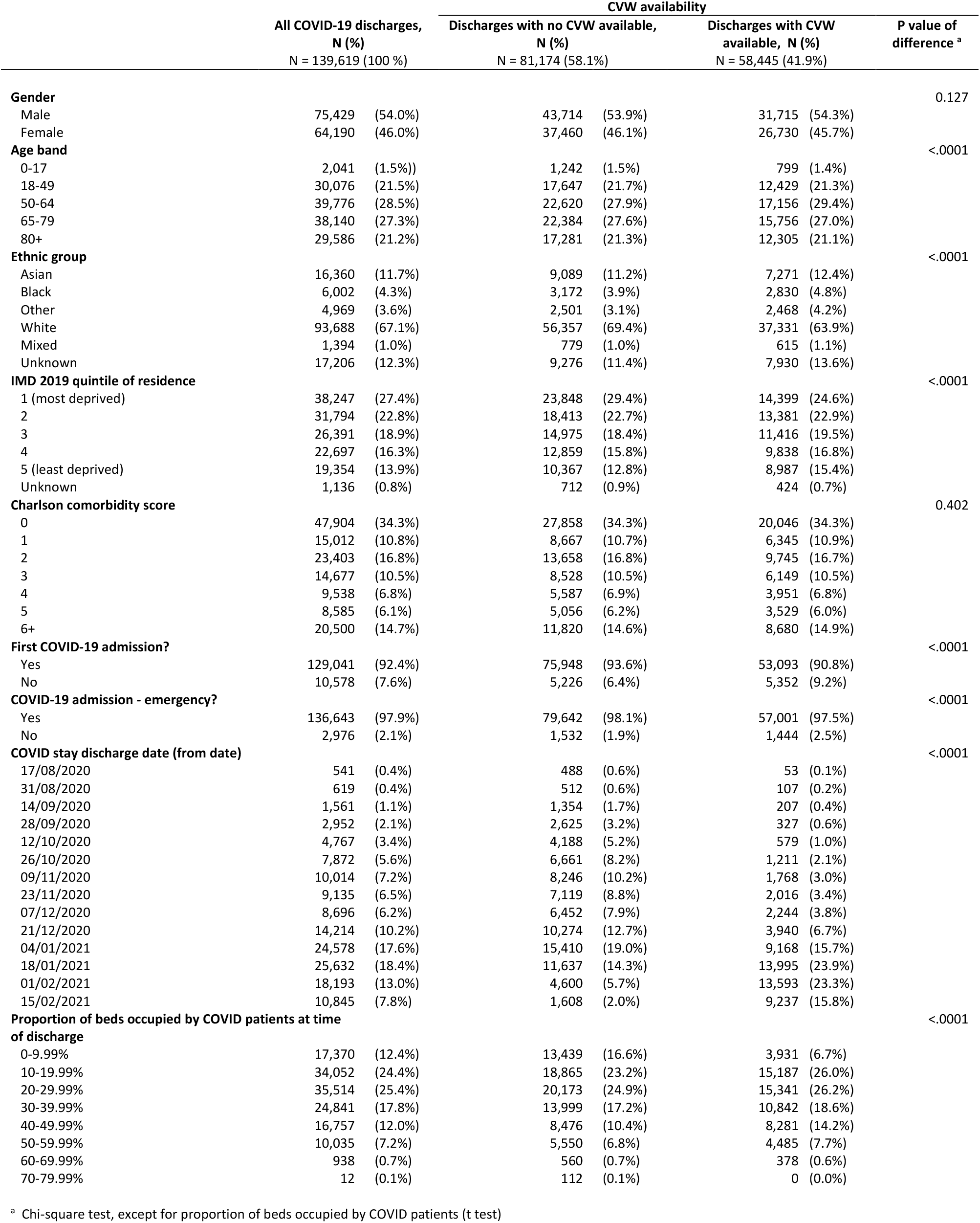
Characteristics of COVID-19 patient discharges, also split by CVW availability.

The number of COVID-19 patients discharged in this time across England (not limited to the 123 analysis trusts) was 142,216. Thus, our analysis includes 98·2% of recorded inpatient stays of patients in England with a primary COVID-19 diagnosis, discharged alive.

Table 3 also shows the characteristics of the patients split into two key analysis groups: patients discharged where no CVW was available (N = 81,174, 58·1%), and those discharged where a CVW was available (N = 58,445, 41·9%). These two sets of patients were well matched in terms of gender and comorbidities. Patients discharged where a CVW was available were more likely to be in the age range 50 to 64 and members of non-White ethnic groups, and less likely to be from the most deprived areas. As a consequence of the accelerated set up of CVW services from early January 2021 (Figure 1), discharge dates of patients where a CVW was available tended to take place toward the end of the analysis period, with nearly 4 in 5 occurring on or after 4 January 2021 (compared to 2 in 5 of the discharges where a CVW was not available). Discharges of patients taking place where a CVW was available were more likely to be occurring in the context of higher levels of beds occupied by COVID-19 patients.

**Figure 1.**
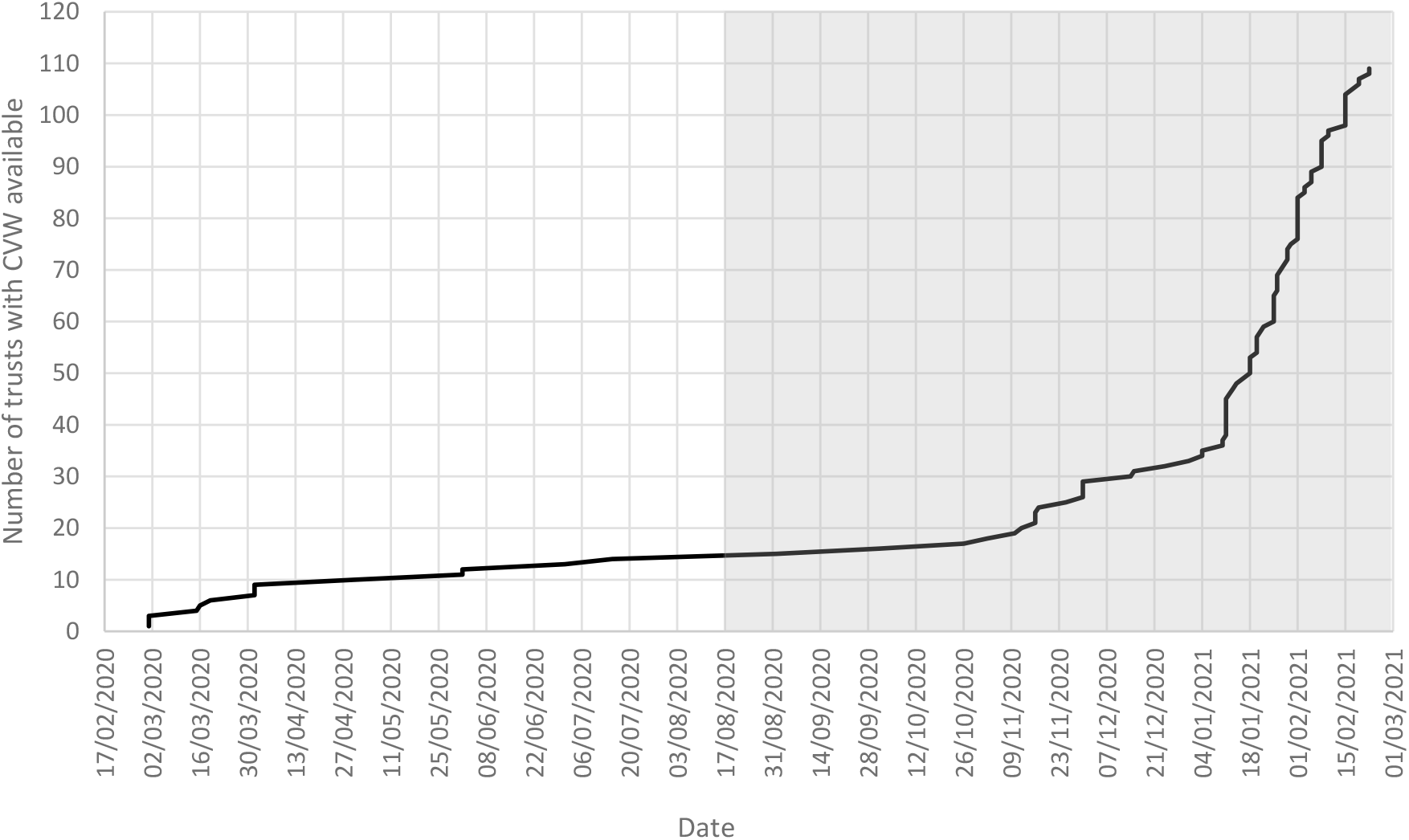
Cumulative start dates of CVW services in England’s non-specialist acute hospital trusts (n = 123 trusts). 14 trusts had no CVW by 22 February 2021. The grey box represents the main period of this analysis.

### COVID-19 length of stay

The mean LOS for patients discharged where a CVW was available was 10·1 days (standard deviation (SD) 11·3 days), compared to 8·9 days (SD 10·3 days) for patients where a CVW was not available (Figure 2); this was an unadjusted incidence rate ratio of 1·13, or 13% longer for discharges taking place from hospital trusts with a CVW. Distributions of COVID-19 length of stays for patients discharged with and without an available CVW are provided in the Supplementary material (Figure S1).

**Figure 2.**
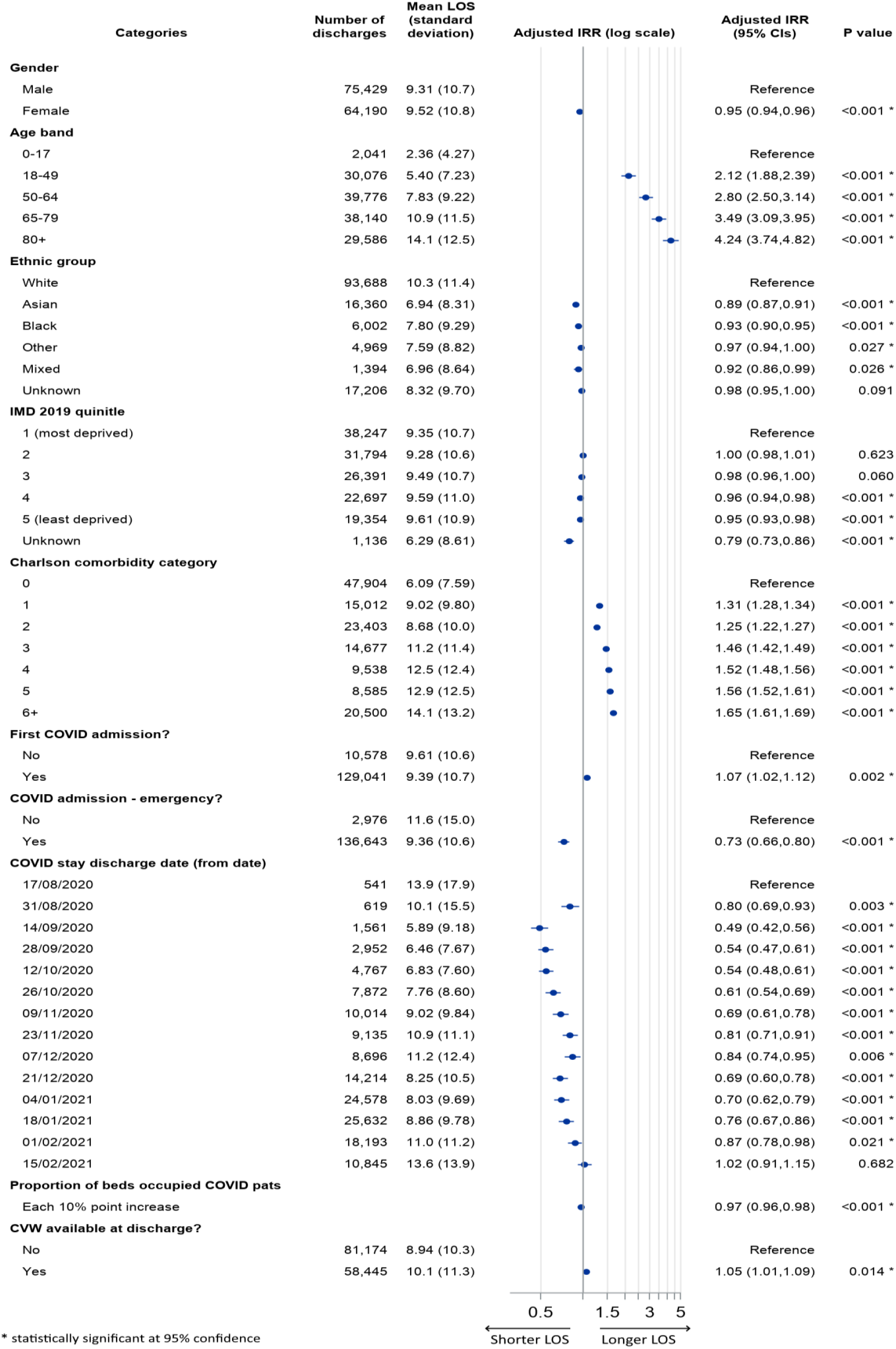
Mean (unadjusted) LOS of COVID-19 hospitalisations, and adjusted Incidence Rate Ratios (IRR), with respect to categories of factors in multivariate model.

Fully adjusting for all covariates, we found an adjusted incidence rate ratio (IRR) of 1·05 (95% CI 1·01 to 1·09) for discharges where a CVW was available compared to those where a CVW was not available, indicating that the length of stay was 5% longer for relevant COVID-19 inpatient stays where a CVW had been available.

In terms of the other factors in the fully adjusted model, we additionally found strong, generally positive gradients of LOS for increasing age and comorbidity (note that these relationships relate to all 139,619 discharges, and not just those with a CVW-available). Females had shorter lengths of stay than males (adjusted IRR 0·95, 95% CI 0·94 to 0·96), as did all non-White ethnic groupings, compared to the White group (for example, Asian patients had an adjusted IRR 0·89, 95% CI 0·87 to 0·91, and Black patients an adjusted IRR of 0·93, 95% CI 0·90 to 0·95). Patients resident in the most affluent population quintiles tended to have slightly shorter lengths of stay than those in the most deprived decile. First COVID-19 inpatient stays were longer than subsequent stays (adjusted IRR 1·07, 95% CI 1·02 to 1·12). The LOS shortened significantly for all time periods after 31 August 2020, until mid-February 2021. As the proportion of acute beds occupied by COVID-19 patients increased, LOS tended to fall: for every ten-percentage point increase in this proportion, the adjusted IRR was 0·97 (CI 0·96 to 0·98).

Model variants carried out as sensitivity analyses marginally altered the resulting adjusted IRR of the CVW-available discharges (Supplementary Table S1). Shortening the time period variable to 7 days reduced the IRR slightly such that the difference was no longer significant at 95% statistical confidence level (IRR 1·04, 95% CI 0·997 to 1·08), while increasing the period to 28 days increased the adjusted IRR to 1·08 (95% CI 1·04 to 1·13). The addition of earlier (wave 1) data made the two groups effectively indistinguishable from one another (IRR 1·01, 95% CI 0·97 to 1·05). The two alternative LOS outcomes tested had little impact on the adjusted IRRs compared to the base model.

### COVID-19 readmissions within 28 days

Of the patients discharged from trusts with a CVW available, 13.0% were readmitted with COVID-19 within 28 days, compared to 13·2% from trusts where no CVW was available (Figure 3). Adjusting for all variables, we found a non-significant difference in COVID-19 readmissions within 28 days, for discharges where a CVW was available compared to those where a CVW was not available (adjusted odds ratio (OR) of 0·97, 95% CI 0·91 to 1·03). Our sensitivity analyses demonstrated the robustness of these findings.

**Figure 3.**
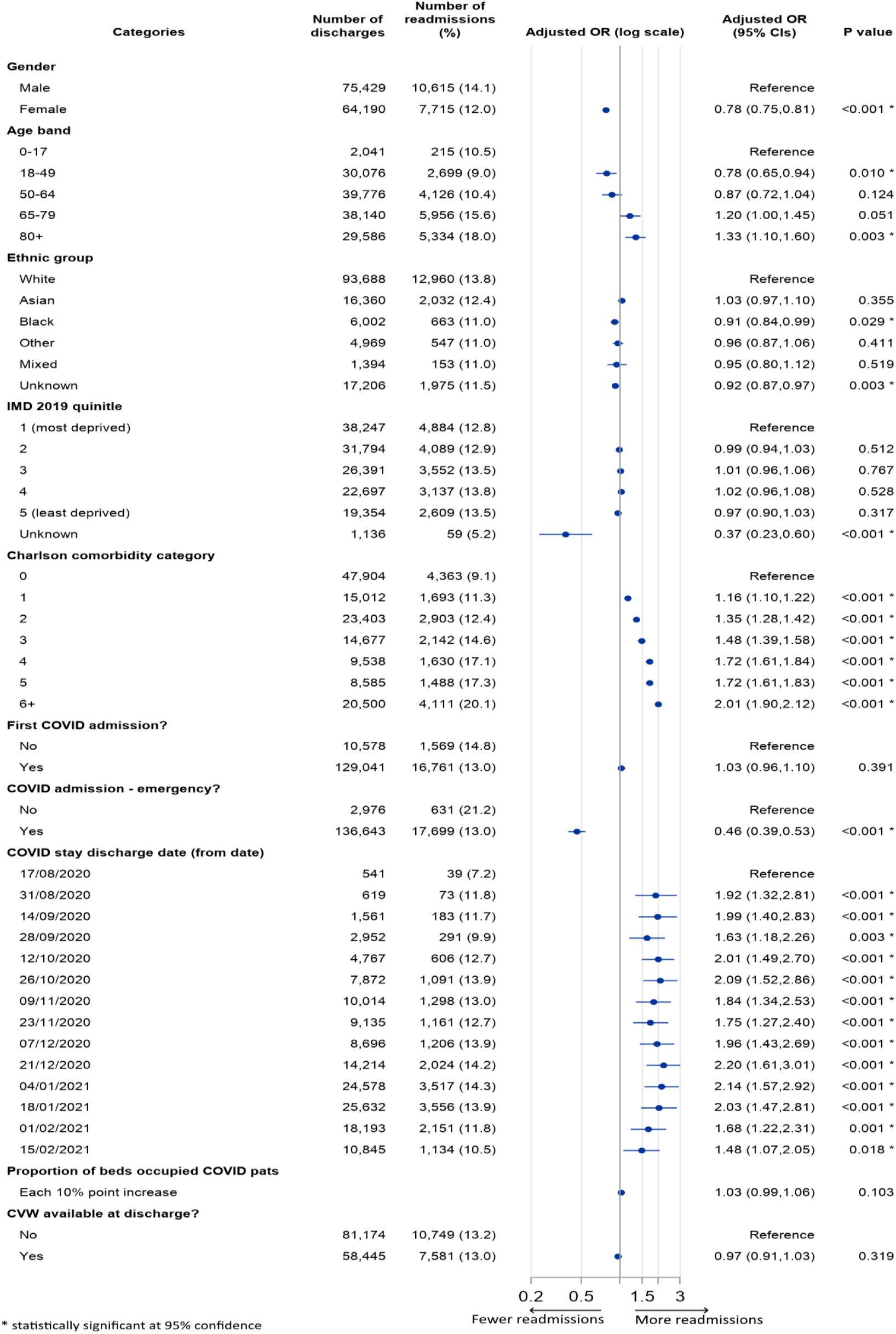
Numbers (%) of patients readmitted for COVID-19 within 28 days, and adjusted Odds Ratios (OR), with respect to factors in multivariate model.

As with LOS, we found generally positive gradients of relative odds of readmission with increasing age and comorbidity. Females had significantly lower odds of readmission than males (adjusted OR 0·78, 95% CI 0·75 to 0·81). Black patients had lower odds of readmission compared to White patients (adjusted OR 0·91, 95% CI 0·84 to 0·99), while no other ethnic categories (for patients with known ethnicity) showed similarly significant differences. There were no apparent differences in the odds of COVID-19 readmission for patients resident in less deprived areas, compared to those in the most deprived quintile areas.

For both LOS and readmissions models, adding variables one by one to our multivariate models (an example shown in supplementary table S2) revealed that the addition of the 14-day time period categories, and the clustering by hospital trust had large impacts on the estimated adjusted ratios, but all additional variables (age, comorbidity index, and so on) had relatively marginal additional impacts.

## Discussion

In this analysis of COVID-19 patients discharged from hospital during the second wave of the COVID-19 pandemic in England, we found no evidence of a relationship between the availability of hospital-based COVID virtual wards (CVW) and subsequent rates of readmission for COVID-19. We did, however, find that the length of stay for hospitalised COVID-19 patients was 5% longer where a CVW was available.

There are few studies available against which we can compare our results. Our finding of no difference with respect to readmissions appears to contrast with that of an analysis of a post-discharge remote monitoring model implemented in five Boston (US) hospitals. This study found that enrolled patients had a reduced odds (OR 0·54, p-value 0·04) of attending ED or being readmitted within 30 days of discharge, compared to those not enrolled.^18^ However, given the relatively large confidence intervals in their estimate, ED attendances being much more common than readmissions for the enrolled patient cohort, and those readmissions being for any cause, our findings may not be inconsistent.

Two other analyses have claimed reductions in length of hospitalisation associated with post-discharge models, on a scale of 30-40%. One, related to implementation in a Netherlands hospital, estimated a reduction in the LOS of 5·0 days, (with a resulting mean LOS of 10·6 days),^19^ and a second, in an English hospital trust reported a reduced LOS of 10 days, against an average of 17 days prior to implementation.^6^ Both analyses, however, were based on small patient cohorts with no formal statistical testing; in the first analysis it was not clear how estimates of reductions were made, and in the second there was no attempt made to control for differences between the compared groups.

More generally, there is evidence of non-COVID related post-discharge virtual wards having a positive impact on readmissions when employed as a disease-specific intervention (at least in the case of heart failure), but there is not similar evidence of impact for more mixed groups of high-risk chronic disease patients.^12^ Our findings may be consistent with this picture, considering the heterogeneity of COVID-19 patients, and the multi-organ effects of COVID-19 infection.^30,37,38^

### Strengths and limitations

With respect to the analysis of post-discharge COVID monitoring services our study is currently unique in its scope. We have made use of a national administrative hospital dataset, and have analysed almost 140,000 COVID-19 admissions in an effort to detect an impact of a national-scale roll out of post-discharge remote monitoring services. We have been pragmatic in our use of the available data, and have controlled for characteristics available to us.

Nevertheless, there are several limitations which mean that our results should be interpreted with caution. We did not know which patients were enrolled onto a CVW, and so treated all patients as potentially having received the intervention where one was available at a hospital trust. We had no information about important clinical factors, for example: ICU admission, clinical readings (including oxygen saturation levels), and specific treatments received. The location to which patients were discharged (home, care home, and so on) was not known, and we also had no information about out-of-hospital deaths (and this during a period in which rates of out-of-hospital deaths had been persistently above long-term norms)^39^. We extracted data only on patients discharged alive, and so the impact (if any) of COVID-19 patients who died during their hospital stay has not been accounted for. Our analyses have assumed that these factors – which are likely to have been associated to a greater or lesser degree with eligibility for referral, and/or with risks of readmission and long stays – were well balanced between the two key groupings of patients after having adjusted for model factors. But the extent to which this is the case is not known. However, in terms of at least one of these missing factors, an analysis of a cohort of 419 COVID-19 patients managed on a post-discharge pathway suggests that out-of-hospital deaths may have been relatively rare (<0·1%).^5^

From our wider study of oximetry monitoring programmes in 28 sites, we had some information about the implementation of CVW services in a limited number of sites, including reports of numbers of patients monitored.^16^ For seven hospital trust-based sites, we estimated a range of from 4% up to 65% of discharged COVID-19 patients may have been monitored on a CVW. But nationally, we did not know the scale of each trust’s implementation and our analysis was not able to examine potential dose-response relationships.

Our study also indicated that services were implemented with significant differences, depending on the locally adopted models of care and whether they were running an integrated CVW/CO@h service: our analysis treated the intervention as being homogeneous, and was not able to estimate differential impacts by hospital trust, though these may have existed.

### Interpretation

We found longer COVID-19 inpatient stays associated with implementation of COVID virtual wards; this was not expected for a service that aimed to support the early discharge of patients. One explanation might be that CVW eligibility criteria influenced hospitals’ discharge decisions such that patients were kept in a bed for longer than they otherwise might have been. It is not clear whether local pre-hospital (CO@h) services starting at similar times might have affected admissions in such a way that patients were admitted sooner, and stayed for longer. The finding might in part also be a consequence of many CVW services launching during the falling (improving) edge of England’s second COVID-19 wave, with longer lengths of stay being a consequence of improving bed capacity.

We did attempt to control for the latter effect in our addition of trust-specific weekly occupied beds data, and with our inclusion of a time period variable in our multivariate analyses. In one sensitivity analysis, when we shortened the time period to 7 days (from 14) – we no longer found a statistically significant difference in LOS at the 95% level (adjusted IRR 1·04, 95% CI 1·00 to 1·08, p-value 0·07). The time factor used in our fully-adjusted models was somewhat arbitrary, and it is not clear that 14 days was better used above 7 days. The finding of a longer LOS associated with CVW therefore needs to be considered with some care.

As a result of the limitations in the available data outlined above, there is a strong possibility that any positive impacts - at a local hospital trust level, or for specific versions of the pathway - were not ultimately able to be detected. This limits the extent to which this analysis can contribute to the further development of these, or similar services, and highlights the importance of making routine data collection - appropriate for use in evaluation – an intrinsic part of services’ operations.

Briefly, we reflect on observed relationships between patient characteristics and COVID-19 lengths of stay, and readmissions. Positive relationships between age and comorbidities and these outcomes have been observed elsewhere.^28,32,40,41^ Meanwhile, differences with respect to gender (in this study’s case both outcomes were lower for females versus males) tend to have been absent.^29,30,32,41^ We found shorter lengths of stay and lower rates of readmissions for patients belonging to specific ethnic groups with respect to White patients (this was the case for patients belonging to Black ethnic groups for both outcomes, and also Asian, Mixed and ‘Other’ ethnic groups for length of stay). Evidence from elsewhere is mixed, but at least one US study found shorter lengths of stay for hospitalised non-White patient groups.^41^ Ultimately, however, care is needed when interpreting findings for specific characteristics separate from the context of the other model variables; it is possible, for example that non-White ethnic groups (to give one example) may be acting as fine-scale markers of urban or more deprived areas (in addition to the model deprivation categories) and as markers of younger patients (within modelled age bands).

### Conclusion

Our analysis has not shown any evidence of early discharges or reduced rates of readmissions associated with the roll out of COVID Virtual Wards in England. While this may reflect the true impact of the service, it may be in part a consequence of the lack of certain data: on which patients were enrolled (or even how many), what COVID-19 treatments were received while in hospital, their clinical observations. It is possible that CVW services had a range of impacts, that we have ultimately not been able to reveal.

## Supporting information

STROBE checklist

## Data Availability

Individual patient-level data and data supplied under specific data sharing agreements cannot be made available by the study team. Sources for data that are already publicly available are supplied either in the text or the references.

https://digital.nhs.uk/services/data-access-request-service-dars

## Acknowledgements

Thank you to the following: our NIHR BRACE and NIHR RSET public patient involvement members; Dr Jennifer Bousfield, Dr Jo Ellins, Dr Ian Litchfield, Jon Sussex for advice given throughout the project, Pei Li Ng for advice and practical support throughout the project, the NIHR 70@70 Senior Nurse research Leaders for providing feedback on the development of our study; Russell Mannion for peer-reviewing our study protocol; the NIHR Clinical Research Networks for supporting study set up and data collection; Public Health England and the Institute of Global Health Innovation, NIHR Patient Safety Translational Research centre, Imperial College London, NHS England and Kent, Surrey and Sussex Academic Health Sciences Network for providing data; Dr Cono Ariti (University of Cardiff) for statistical advice; and Eilís Keeble (Nuffield Trust) for her approach to classifying COVID-19 spells in Hospital Episode Statistics.

We thank the NHS Digital CO@h Evaluation Workstream Group chaired by Professor Jonathan Benger for facilitating and supporting the evaluation, and to the other two evaluation teams for their collaboration throughout this evaluation: i) Institute of Global Health Innovation, NIHR Patient Safety Translational Research centre, Imperial College London and ii) the Improvement Analytics Unit (Partnership between the Health Foundation and NHS England and NHS Improvement).

Many thanks to our Clinical Advisory Group for providing insights and feedback throughout the project (Dr Karen Kirkham (whose previous role was the Integrated Care System Clinical Lead, NHSE/I Senior Medical Advisor Primary Care Transformation, Senior Medical Advisor to the Primary Care Provider Transformation team), Dr Matt Inada-Kim (Clinical Lead Deterioration & National Specialist Advisor Sepsis, National Clinical Lead - Deterioration & Specialist Advisor Deterioration, NHS England & Improvement) and Dr Allison Streetly (Senior Public Health Advisor, Deputy National Lead, Healthcare Public Health, Medical Directorate NHS England).

Hospital Episode Statistics data are reused with permission of NHS Digital (Copyright © 2022, NHS Digital. All rights reserved).

## Contributors’ statement

TG, CSJ, SM, EM and SMT contributed to study design and methodology. TG and CSJ participated in data curation, analysis and validation. TG and CSJ have verified the underlying data. NJF led the overall mixed methods evaluation of which this study relates to one workstream. All authors provided input from their own workstreams to help interpret findings. TG led on the preparation and writing of the manuscript. All authors reviewed and provided feedback on the manuscript and approved the final version.

