## Supplementary Material for "The impact of post-hospital remote monitoring of COVID-19 patients using pulse oximetry: a national observational study using hospital activity data"

**Contents**


| | | |
|---|---|---|
| Figure S1. | Length of stay distribution for included Covid-19 spells. | Page 2 |
| Table S1. | Sensitivity analyses: variants of analytical models. | Page 2 |
| Table S2. | Sensitivity analyses 2: effect of adding adjustment variables in turn | Page 3 |

**Figure S1. Length of stay distribution for included Covid-19 spells, split by whether CVW was available or not; proportion (a) and cumulative proportions (b) of each group. Of the discharges where no CVW was available 0.79% had LOS > 60 days. For discharges with CVW available 0.92% had LOS > 60 days. All such discharges were set to 60 days in our models.**

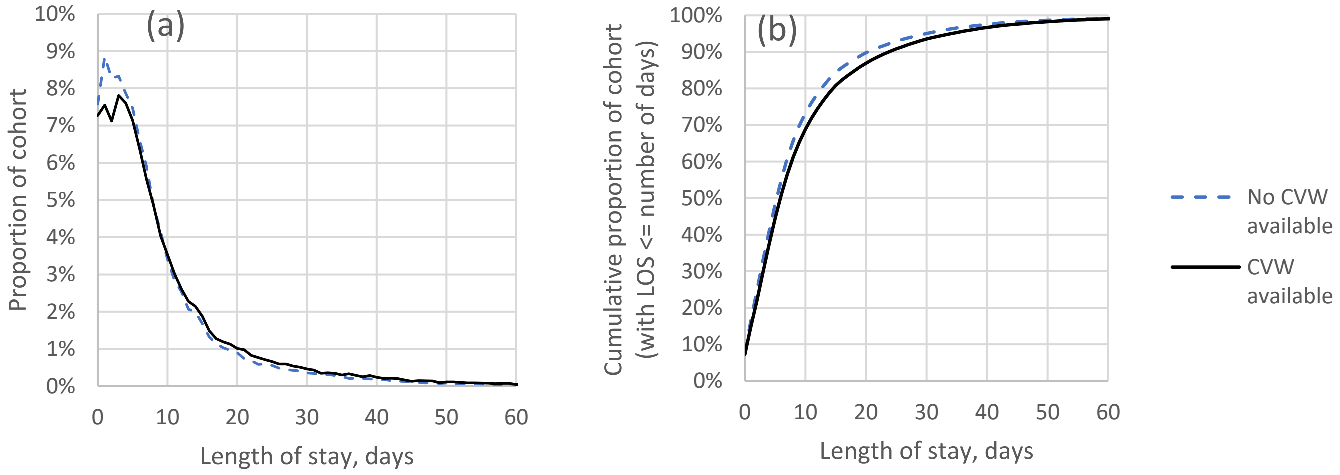


**Table S1. Sensitivity analyses: variants of analytical models. Adjusted Incidence Rate Ratios (IRR) for LOS of COVID-19 spell, and adjusted Odds Ratios (OR) of COVID-19 readmissions within 28 days: discharges with CVW available, vs not available.**

| | **Length of stay of COVID-19 spell** | | | | **Readmission for COVID-19 within 28 days** | | | |
|---|---|---|---|---|---|---|---|---|
| | **IRR** | **P value** | **IRR 95% confidence interval** | | **OR** | **P value** | **OR 95% confidence interval** | |
| | | | **Lower** | **Upper** | | | **Lower** | **Upper** |
| Base models (figs 2 and 3) | 1.05 | 0.01 | 1.01 | 1.09 | 0.97 | 0.32 | 0.91 | 1.03 |
| Time period: 7 days | 1.04 | 0.07 | 1.00 | 1.08 | 0.98 | 0.44 | 0.92 | 1.04 |
| Time period: 28 days | 1.08 | 0.00 | 1.04 | 1.12 | 0.95 | 0.14 | 0.90 | 1.02 |
| Add Wave 1 data | 1.01 | 0.62 | 0.97 | 1.05 | 0.99 | 0.76 | 0.92 | 1.07 |
| LOS untrimmed at 60 days | 1.04 | 0.04 | 1.00 | 1.08 | - | - | - | - |
| LOS disregarding pre-Covid diagnosis | 1.05 | 0.00 | 1.02 | 1.09 | - | - | - | - |

**Table S2. Sensitivity analyses 2: Effect of adding adjustment variables in turn. Adjusted Incidence Rate Ratios (IRR) for LOS of COVID-19 spell, and adjusted Odds Ratios (OR) of COVID-19 readmissions within 28 days: discharges with CVW available, vs not available. Only one ordering shown.**

| | Length of stay of COVID-19 spell | | | | Readmission for COVID-19 within 28 days | | | |
|---|---|---|---|---|---|---|---|---|
| | IRR | P value | IRR 95% confidence interval | | OR | P value | OR 95% confidence interval | |
| | | | Lower | Upper | | | Lower | Upper |
| Unadjusted | 1.13 | <.0001 | 1.11 | 1.14 | 0.98 | 0.14 | 0.95 | 1.01 |
| Adjusted: By time period | 1.00 | 0.88 | 0.99 | 1.01 | 1.03 | 0.14 | 0.99 | 1.06 |
| + (Cluster by provider trust) | 1.05 | 0.03 | 1.00 | 1.10 | 0.98 | 0.50 | 0.92 | 1.04 |
| + Age | 1.05 | 0.02 | 1.01 | 1.09 | 0.99 | 0.67 | 0.93 | 1.05 |
| + Charlson | 1.05 | 0.03 | 1.01 | 1.09 | 0.98 | 0.56 | 0.92 | 1.05 |
| + Deprivation +Ethnic group +Gender | 1.05 | 0.02 | 1.01 | 1.09 | 0.98 | 0.47 | 0.92 | 1.04 |
| + Emergency +First +Proportion of beds occupied COVID | 1.05 | 0.01 | 1.01 | 1.09 | 0.97 | 0.32 | 0.91 | 1.03 |
